# Aesthetic surgeries in body dysmorphic disorder: clinical and gender-specific patterns

**DOI:** 10.1101/2025.11.17.25340436

**Authors:** Judith Diele, Anna Witte, Jens Barenbrügge, Johanna Schulte, Ulrike Buhlmann

## Abstract

Body dysmorphic disorder (BDD) is characterized by an excessive preoccupation with perceived appearance flaws, which is frequently associated with high psychological burden and engagement in cosmetic procedures (instead of seeking psychological help). This study exploratively examined clinical characteristics and the utilization of cosmetic procedures in a large online sample (*N* = 3,240), including – based on self-report – individuals with BDD (*n* = 1,837), subclinical symptoms (*n* = 392), and a control group with appearance concerns (*n* = 166). Participants completed standardized self-report measures assessing BDD symptoms, appearance concerns, mental health history, and cosmetic surgery experiences. Results indicated that approximately 30 % of individuals meeting full DSM-5 criteria for BDD had undergone at least one cosmetic procedure, with an even larger proportion interested in future surgeries. Satisfaction with past interventions was rather low, supporting prior evidence that cosmetic treatments rarely address the underlying psychopathology of BDD. Hierarchical regression analyses yielded that both BDD symptom severity and lower illness insight significantly predicted cosmetic surgery intent, whereas only symptom severity, but not insight, was associated with past surgery behavior.

Furthermore, gender-based analyses revealed that while women and men did not differ in overall surgery rates, women more often pursued or planned facial and breast-related procedures, whereas men more frequently reported hair-related concerns and interventions. Women (vs. men) also reported higher symptom severity and greater barriers to accessing surgery, such as financial concerns and discouragement from people in their personal lives. Notably, more than 90 % of individuals with BDD had never received a formal BDD diagnosis, underscoring treatment barriers and under-recognition of BDD in clinical settings. These findings highlight the need for early screening, especially in cosmetic and dermatological contexts, as well as treatment concepts that include specific modules addressing the wish for cosmetic procedures.

## Introduction

Body dysmorphic disorder (BDD) is characterized by an extensive preoccupation with one or more perceived appearance flaws that are not or only slightly visible to others [1]. This preoccupation causes significant distress and impairment in social, occupational, and academic functioning, resulting in a reduced quality of life [e.g., 2]. Individuals with BDD frequently engage in excessive or repetitive appearance-related behaviors, such as mirror checking, camouflaging perceived flaws, or reassurance seeking, as well as mental acts like comparing one’s own appearance to that of others [1]. The estimated point prevalence of BDD in the general adult population is approximately 1.9 % [e.g., 3,4], a rate that surpasses that of other severe mental disorders such as anorexia nervosa and schizophrenia [1].

Despite its high prevalence, BDD often goes underdiagnosed and untreated. A key factor contributing to this problem is the appearance-related shame, which often leads affected individuals to conceal their symptoms [5,6]. Moreover, many individuals with BDD have limited insight into the psychological origins of their distress and firmly believe that only cosmetic or medical interventions can alleviate their perceived flaws [7]. Insight in BDD is not static but can also vary over time and across situations, which might further complicate clinical identification [8]. Thus, both shame and limited insight can contribute to a significant delay in seeking psychological or psychiatric treatment, sometimes for years, while individuals attempt to correct their appearance through aesthetic procedures [3,6]. However, such interventions rarely improve symptoms and may even exacerbate the preoccupation with appearance [9,10]. The high rates of suicidality associated with BDD [11] further highlight the need for early identification and a better understanding of how many individuals affected seek aesthetic surgeries instead of psychological or psychiatric help.

Existing studies on cosmetic treatment in BDD often focus on either clinical or non-BDD populations, without adequately comparing subclinical individuals. Online-based study designs offer a valuable alternative by lowering access barriers and enabling participation from individuals who might otherwise be unreachable, including those with subclinical symptoms or who avoid psychotherapeutic treatment altogether. This can enhance the ecological validity and generalizability of findings [3].

Despite the growing cultural emphasis on physical appearance and the rapid increase in aesthetic procedures worldwide [12,13], research exploring the relationship between the utilization of cosmetic treatments and body image disorders such as BDD remains limited [3]. Prevalence estimates of aesthetic surgery among individuals with BDD vary considerably depending on sample characteristics and procedure types, ranging from approximately 13 % to over 50 % [3]. In contrast, in the general population, 3.6 % of adults in Germany and 17.7 % in the U.S. underwent cosmetic surgery in 2023 [13]. Thus, while lower-end estimates in BDD samples can be similar to general population rates, most studies indicate markedly higher prevalence among individuals with BDD [3].

While BDD affects both men and women at comparable rates [e.g., 4,14], gender-specific analyses remain scarce and often rely on small samples, which limits the generalizability of findings. For example, Phillips et al. [14] found that men and women with BDD undergo cosmetic surgery at similar rates, although differences in body areas of concern were noted. In contrast, Malcom et al. [15] investigated cosmetic surgical outcomes in individuals with BDD but did not address potential gender differences, underscoring a notable gap in the literature.

To address these significant gaps, the present study is one of the first large-scale investigations to systematically examine the prevalence of aesthetic surgery and related characteristics across clinical, subclinical, and asymptomatic BDD symptom groups, with a particular focus on gender-specific differences. Using validated self-report measures in a large, anonymous online sample, we aim to overcome limitations of prior studies.

Specifically, our objectives are to (1) assess the lifetime prevalence of aesthetic surgery across BDD symptom groups, and examine whether illness insight shows an additional association with interest in or history of cosmetic procedures beyond overall symptom severity, and (2) examine gender-specific differences in BDD symptoms and experiences with cosmetic surgery, including frequency, motives, and satisfaction. This approach aims to deepen the understanding of cosmetic treatment utilization and attitudes in BDD, providing insights that can help guiding gender-sensitive considerations in both psychiatric and aesthetic medical contexts and ultimately support improved clinical care and outcomes for individuals with BDD.

## Method

### Subjects

Between March 8^th^ 2019 and February 3^rd^ 2025, a total of 19,522 individuals took part in a well-established online self-test for BDD (called scanBB) at the University of Münster. Participants were excluded for non-completion (*n* = 8,706), lack of consent for data use (*n* = 2,796), repeated participation (*n* = 257) or insufficient language skills (*n* = 1). Additionally, participants who scored ≥ 2 on the SCOFF questionnaire (*n* = 4,552) [16] were excluded to reduce the likelihood of confounding with possible eating disorders, as the SCOFF is a screening tool for core symptoms of anorexia and bulimia nervosa. In total, the sample comprised 3,240 participants (75.65 % female, *n* = 2,451), who were on average 31.94 years old (*SD* = 11.61, range = 18 – 77). The data analyzed in this study were also partly used in the study by Vogel et al. [under review].

Based on self-reports, DSM-5 criteria for BDD were assessed using the BDD-5 [17]. In total, 1,837 participants met DSM-5 criteria for BDD and scored above 23 in the Body Dysmorphic Symptoms Inventory (FKS) [18]. An additional 392 participants presented subclinical symptoms, with FKS scores between 14 and 23 and meeting DSM-5 criteria for BDD, except for criterion C (distress or impairment). Lastly, 166 participants were assigned to an appearance concerns control group, characterized by FKS scores below 14 and not meeting the DSM–5 C criterion. Importantly, fulfillment of other DSM-5 criteria (A or B) was not required in this group. The three subgroups comprised 2,395 individuals. The remaining 845 participants did not meet criteria for any group based on the BDD-5.

### Material

The demographic information was collected, including variables such as age, gender, place of residence (country), highest educational qualification, current occupation, and marital status (see S1 Table).

#### Assessment of BDD symptoms, insight and comorbid symptomology

To evaluate DSM-5 criteria for BDD, the BDD-5 measure [17] was utilized, comprising six dichotomous items that operationalize the DSM-5 criteria. If participants met criteria A, B1 and/or B2, C1 and/or C2, and excluded concerns related to body weight (criterion D), the diagnosis was considered fulfilled [1].

The FKS is a self-report measure to assess BDD symptom severity over the past week. It consists of 18 items, with items 2 and 3 addressing the specific body areas of concern and differentiating BDD from eating disorders. A total score is calculated based on the remaining items. Ratings are made on a 5-point Likert scale ranging from 0 (not at all / never / do not think about it) to 4 (very strongly / more than five times / more than 8 hours per day), yielding a maximum sum score of 64 and a cut-off sum score of 14 indicating potential BDD. Scores above 23 indicate a pronounced level of BDD symptomatology [19]. The FKS has demonstrated high internal consistency of α = .88, sensitivity of .87, and specificity of .93 [18].

Illness insight was assessed using item 10 of the FKS, which captures how much individuals are convinced that certain body parts are ugly. Greater conviction, indicated by higher values, reflects stronger belief in the perceived flaw, indicating lower insight.

To screen for potential comorbid eating disorders, participants completed the SCOFF questionnaire [16], a five-item dichotomous (yes/no) screening tool assessing core symptoms of disordered eating. A score of ≥ 2 indicates a likely presence of eating disorder pathology. The SCOFF has shown good diagnostic accuracy, with meta-analytic evidence supporting its high sensitivity and specificity across translations and populations [20].

Depressive symptoms were assessed using the German version of the Patient Health Questionnaire (PHQ-9) [21,22]. The nine items reflect DSM-5 criteria for major depressive disorder and refer to the past two weeks. Responses are given on a 4-point scale (0 = “not at all” to 3 = “nearly every day”). A sum score of ≥10 indicates clinically relevant symptomatology and has shown good diagnostic accuracy, with high sensitivity and specificity [23].

Additionally, participants’ previous psychiatric diagnoses were assessed via self-report. For descriptive purposes, diagnoses were summarized by diagnosis type as well as dichotomized as any prior diagnosis (yes/no), depending on the analysis.

#### Assessment of cosmetic surgery behavior

In item 16 of the FKS, participants were asked whether they had undergone cosmetic surgical procedures to alter body parts and, if so, how often, with responses given in predefined frequency categories (never, once, twice, three to five times, more than five times). If they responded affirmatively to this item, they were prompted to specify the body areas involved, with multiple selections allowed from a predefined list of 40 body areas, including an option for “other”. Satisfaction with the surgical outcome was rated on a 5-point Likert scale ranging from 0 = “very dissatisfied” to 4 = “very satisfied”. All participants were also asked about desired future cosmetic surgeries, with response options including “yes, already planned”, “yes, but not yet planned” and “no”. Participants who indicated planned or desired future surgeries were asked again to specify body areas using the same list of 40 body regions, allowing multiple selections. Participants who indicated that no future surgeries were planned were further asked to provide reasons for not pursuing (or refraining from further) cosmetic surgeries.

### Procedure

Data were collected though our online self-test for BDD (scanBDD), which includes programmed feedback on the extent of appearance concerns and information about BDD and available treatment options using Unipark software [24]. At the beginning and end of the survey, participants were informed about confidentiality, the voluntary nature of participation, and provided with informed consent options. All questions were mandatory, and programmed filter questions guided participants through relevant sections (e.g., past or planned cosmetic surgeries) to ensure complete data. The feedback also provided information on possible treatment options, with contact details for treatment centers. The study was approved by the Ethics Committee of the Institute of Psychology at the University Münster. The average completion time was 16 minutes (*SD* = 24 minutes).

### Statistical Analyses

All analyses were conducted using R (Version 4.5.0) [25], primarily using the packages tidyverse [26], car [27], effectsize [28], emmeans [29], lsr [30], MASS [31], brant [32], rcompanion [33], and psych [34]. Descriptive analyses were performed to report sample characteristics, as well as data on cosmetic surgeries. Means, standard deviations, and frequencies were calculated for the BDD group, the subclinical group and the appearance concerns control group.

In preparation for the analyses, conducted cosmetic surgeries were dichotomized to indicate whether participants had ever undergone at least one cosmetic procedure (0 = none, 1 = at least one). This dichotomous variable was used for group comparisons. In contrast, for regression analyses within the BDD group, the original count variable indicating the number of procedures was retained and modeled as an ordinal outcome. Moreover, planned and desired future surgeries were combined into a single category to reflect general interest in cosmetic procedures.

First, group comparisons were conducted. Since age was unequally distributed across the groups, linear or logistic regression models were employed to statistically control for age, depending on whether the dependent variable was continuous or categorical. Post-hoc pairwise comparisons between all groups were conducted, irrespective of the statistical significance of the overall group effect, to provide a comprehensive overview of potential group differences. Within the BDD group, separate analyses were also conducted for female and male participants. Gender differences in clinical and demographic variables were examined using independent samples *t*-tests for continuous variables and chi-square tests for categorical variables. In cases where potential confounding by symptom severity was expected, logistic regression models were employed to control for symptom severity when analyzing binary outcomes related to gender differences.

To examine whether conviction was additionally associated with interest in or history of cosmetic procedures beyond symptom severity, two sets of hierarchical regression analyses were conducted. First, ordinal logistic regression was used to predict the number of cosmetic procedures performed. Model 1 included the FKS total score as a predictor excluding item 10 (assessing conviction regarding the perceived defect) and item 16 (assessing history of conducted cosmetic surgeries) to avoid overlap with the outcome variables. Model 2 added conviction (FKS item 10) to evaluate incremental predictive value. Model fit was evaluated using likelihood-ratio tests and Akaike’s Information Criterion (AIC). Second, binary logistic regression was used to predict the desire for cosmetic surgery. Again, Model 1 included the FKS total score (excluding items 10 and 16) as the sole predictor. Model 2 additionally added conviction. Model comparisons were based on likelihood-ratio tests and Nagelkerke’s R².

Effect sizes are reported using Cohen’s *d* for *t*-tests, Cramér’s *V* for chi-square tests, η² for linear regression and odds ratios (*OR*) for logistic regression. An alpha level of .05 was applied to all statistical tests. To control the false discovery rate, *p*-values were adjusted using the Benjamini-Hochberg procedure.

## Results

### Cosmetic surgery and psychopathology across BDD severity groups

A total of 1,837 participants met – based on self-report – DSM-5 criteria for BDD, with ages ranging from 18 to 73 years (*M* = 30.76, *SD* = 10.65). Clinical and demographic characteristics, including appearance concerns, cosmetic surgeries, reasons against cosmetic surgeries, and clinical diagnoses for the total group, are reported in the Supplementary Materials (S1 Table).

To contextualize these findings, comparisons were conducted between the BDD, subclinical BDD, and appearance concerns control groups, controlling for age due to a significantly unequal age distribution, *p* < .001, η² *=* .05. Key clinical differences are summarized in Table 1, with full statistical results available in Supplementary S2 Table.

**Table 1.** Key Clinical and Behavioral Differences between BDD Status Groups.

|  | BDD<br>(*n* = 1,837) | Subclinical BDD<br>(*n* = 392) | Appearance<br>Concerns<br>(*n* = 166) | Comparison |  |  |
| --- | --- | --- | --- | --- | --- | --- |
|  | *M* (*SD*) / %<br>(*n*) | *M* (*SD*) / % (*n*) | *M* (*SD*) / %<br>(*n*) | BDD vs. Subclinical<br>BDD<br>*p*<sub>adj</sub> (*OR*) | BDD vs.<br>Appearance<br>Concerns<br>*p*<sub>adj</sub> (*OR*) | Appearance<br>Concerns vs.<br>Subclinical BDD<br>*p*<sub>adj</sub> (*OR*) |
| College/university degree | 35.11 (645) | 46.17 (181) | 56.63 (94) | .008** (1.41) | .006** (1.70) | .412 (0.83) |
| Clinical factors |  |  |  |  |  |  |
| PHQ-9 Score | 12.39 (5.86) | 6.86 (4.45) | 4.17 (4.07) | < .001*** (0.20<sup>†</sup>) | < .001*** (0.20<sup>†</sup>) | < .001*** (0.20<sup>†</sup>) |
| Prior Clinical Diagnoses |  |  |  |  |  |  |
| Affective Disorder | 45.51 (836) | 31.63 (124) | 26.51 (44) | < .001*** (2.06) | < .001*** (3.28) | .052 (0.63) |
| Social Anxiety Disorder | 15.90 (292) | 8.42 (33) | 3.01 (5) | < .001*** (2.03) | < .001*** (5.88) | .054 (0.35) |
| Personality Disorder | 12.47 (229) | 6.89 (27) | 1.81 (3) | .011* (2.10) | < .001*** (9.77) | .026* (0.22) |
| BDD | 9.42 (173) | 1.28 (5) | 1.20 (2) | < .001*** (9.31) | .011* (12.17) | .773 (0.77) |
| Cosmetic Surgery Behavior |  |  |  |  |  |  |
| Conducted Cosmetic<br>Surgery | 29.50 (542) | 15.82 (62) | 17.47 (29) | < .001*** (0.38) | < .001*** (0.15) | .007** (2.60) |
| Satisfaction with Cosmetic<br>Surgery | 1.89 (1.27) | 2.22 (1.26) | 2.93 (1.21) | < .001*** (0.02<sup>†</sup>) | < .001*** (0.02<sup>†</sup>) | < .001*** (0.02<sup>†</sup>) |
| Future Cosmetic Surgery | 68.97 (1,262) | 39.54 (155) | 15.05 (25) | < .001*** (3.30) | < .001*** (12.10) | < .001*** (0.28) |
| Reasons against Surgery:<br>Fear of looking worse | 40.94 (411) | 28.15 (38) | 20.00 (4) | .011* (1.77) | .122 (2.77) | .518 (0.64) |

BDD = Body Dysmorphic Disorder. <sup>†</sup> Partial eta-squared ( $\eta^2$ ). \*\*\**p* < .001; \*\**p* < .01; \**p* < .05

As expected, the BDD (vs. subclinical and appearance concerns control) group exhibited higher average BDD symptom severity and significantly more depressive symptom burden. They were also more likely to report prior diagnoses of affective, social anxiety, and personality disorders and less likely to report no mental health history. While nearly half of the BDD group (44.52 %, *n* = 818) reported having received one or more professional diagnoses, only a small minority had received a formal BDD diagnosis. Additionally, educational differences emerged: even after controlling for age, individuals with BDD were significantly less likely to hold a university degree than the remaining groups.

In terms of cosmetic surgery behavior, individuals with BDD were more likely to have undergone at least one cosmetic surgery than the other groups. Despite this elevated frequency, satisfaction with surgical outcomes was notably lower. Moreover, the prevalence of desired cosmetic surgeries was significantly higher among individuals with BDD who had previously undergone cosmetic surgery, *p* < .001, *V* = 0.29.

Interest in future cosmetic procedures was substantially higher in the BDD group, with more than two thirds of the participants expressing either concrete plans or general intent. Planned or desired surgeries most frequently targeted the nose, followed by the face and breast, corresponding with the most commonly performed surgeries in the BDD group. However, this only partially overlaps with reported appearance concerns, which most frequently centered face, nose, and skin. Regarding reasons against undergoing cosmetic procedures, financial constraints were the most commonly cited barrier across all groups. While additional reasons were also reported, only the group difference for fear of looking worse reached statistical significance (significant for BDD vs. subclinical BDD, not for BDD vs. appearance concerns control group).

Additionally, two hierarchical logistic regression models examined whether conviction showed an additional association with interest in or history of cosmetic procedures beyond symptom severity (FKS total score, excluding items 10 and 16 for comparability). Regarding past cosmetic surgeries, higher symptom severity was significantly associated with having undergone surgery, *B* = 0.029, *p* = .009, AIC = 1312.76. Adding conviction did not improve model fit, ΔAIC = 1.01, *p* = .320, and conviction was not a significant predictor, *B* = -0.123, *p* = .325, suggesting that insight did not provide additional explanatory value. In contrast, for interest in future cosmetic surgery, higher BDD symptom severity significantly predicted the likelihood of surgery interest, *OR* = 1.06, *p* < .001, R² = .042. Adding conviction as a predictor significantly improved model fit, ΔR² = .028; *OR* = 1.53, *p* < .001, indicating both symptom severity and lower insight independently contributed to surgery interest.

### Gender differences in individuals with BDD

The key gender-specific differences in the BDD group are provided in Table 2 (for full results, see Supplementary S3 Table). Women reported significantly higher BDD symptom severity as measured by the FKS. Therefore, several group comparisons were statistically adjusted for symptom severity to ensure that observed differences were not confounded by differences in symptom burden.

**Table 2.**
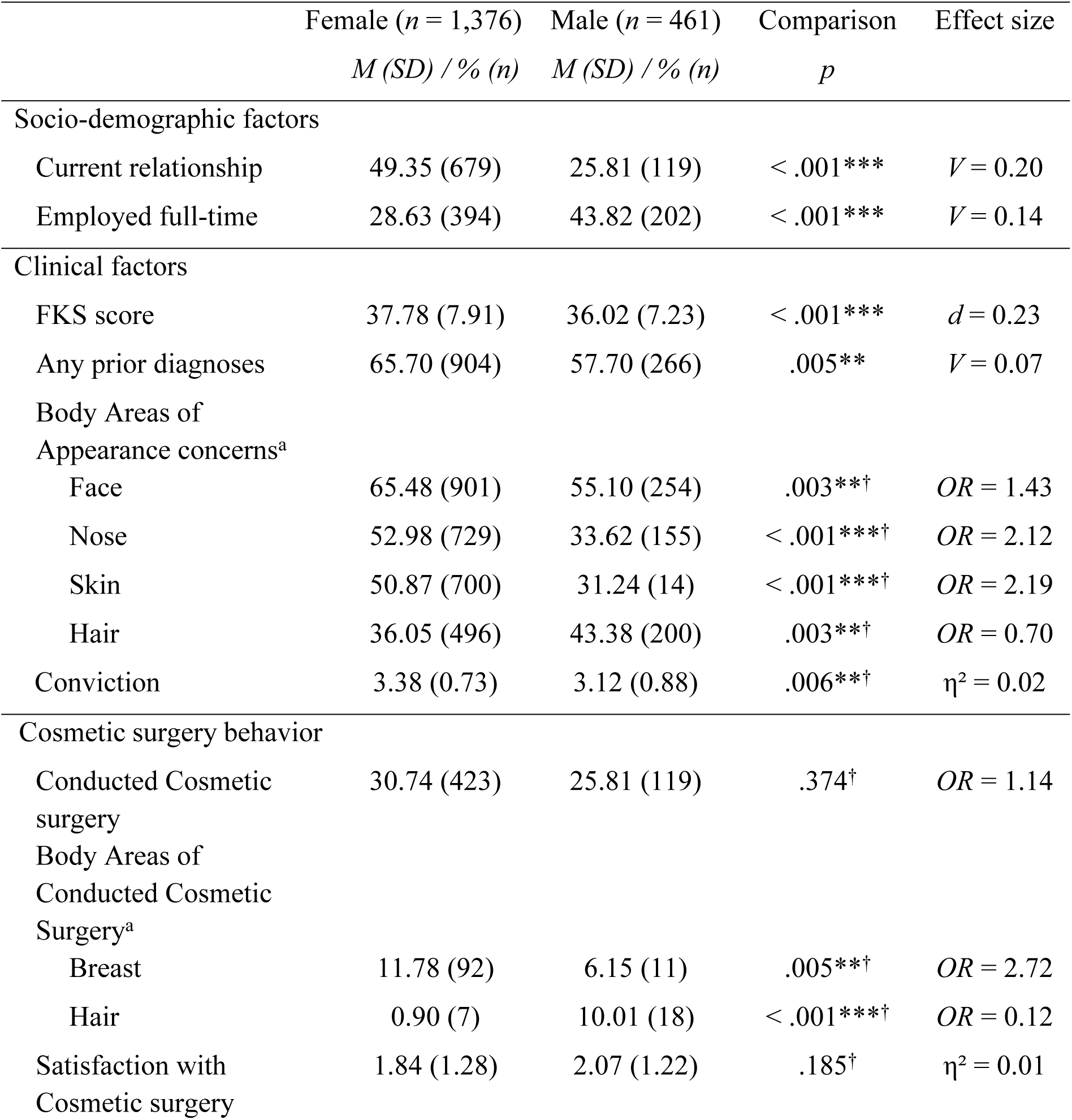
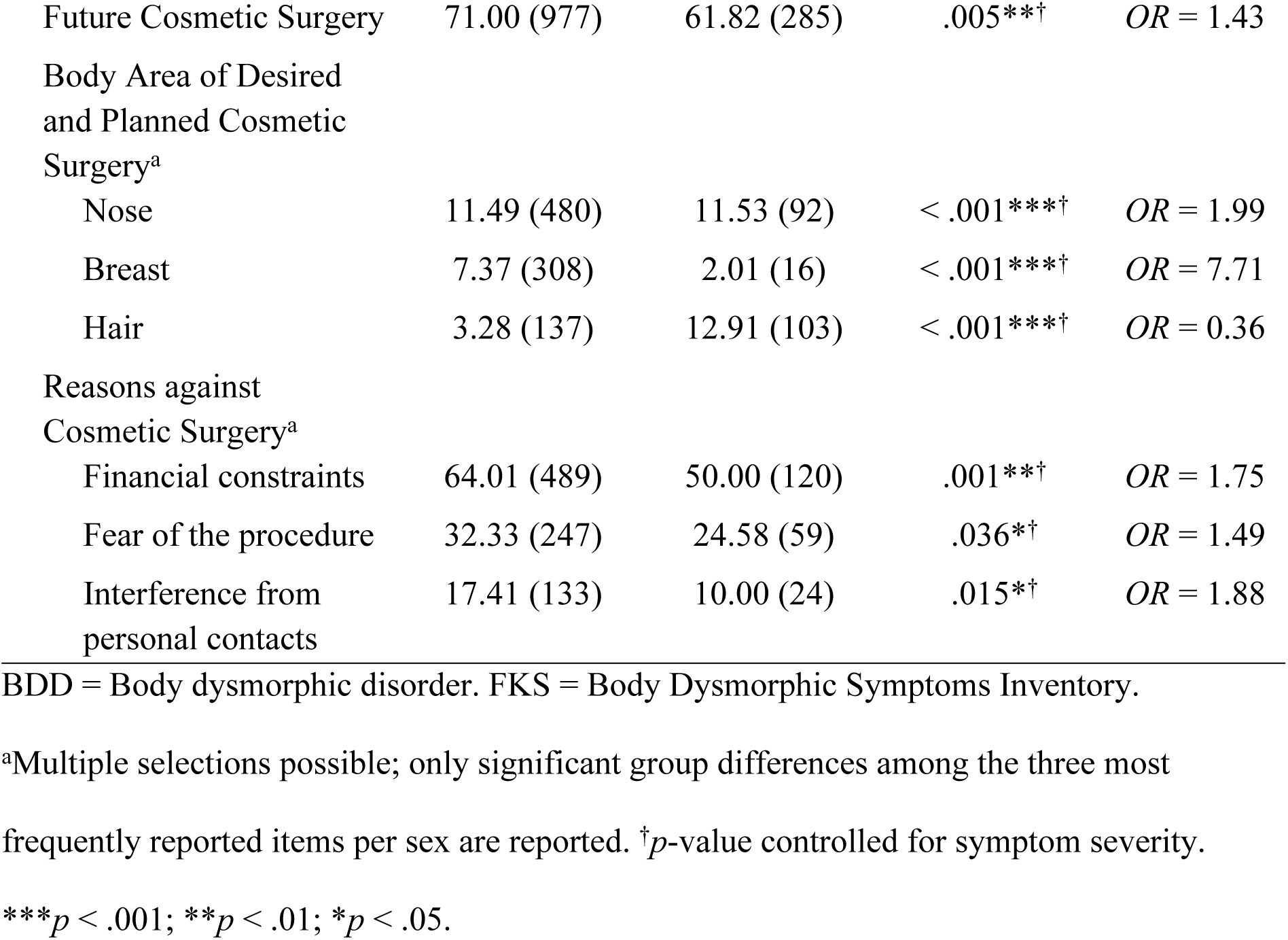
Key Gender Differences in Sociodemographic, Clinical, and Cosmetic Surgery Variables among Individuals with BDD.

Sociodemographic and clinical differences revealed that women were more likely to be in a current relationship, whereas men were more often employed full-time. No significant gender differences emerged in educational level or age. Regarding clinical diagnoses, men were significantly more likely to have never received a prior clinical diagnosis, while no gender differences emerged in the prevalence of affective, social anxiety, personality, or BDD diagnoses.

While the overall prevalence of cosmetic surgery did not significantly differ between women and men, gender-specific patterns emerged in the type of procedures: women were more likely to have undergone breast surgery, whereas hair surgery was more common among men. No significant gender differences were found in the frequency of nose or facial surgeries. Satisfaction with cosmetic procedures was similar across genders. Similarly, within the subgroup of individuals who had undergone cosmetic surgery, no significant gender difference in BDD symptom severity was observed, *p* = .286, *d* = 0.11. However, a significant gender difference emerged in illness insight, with women reporting higher conviction in their appearance-related beliefs than men. Women were also more inclined to express interest in future cosmetic procedures and were more likely to express a wish for breast surgery, whereas men were significantly more interested in hair procedures. When controlling for BDD symptom severity, women additionally showed almost twice the odds of expressing interest in nasal surgery compared to men. In terms of body areas of appearance concern, women were more likely to report distress about their face, nose, and skin while hair-related concerns were more common among men. Among reasons against cosmetic surgery, women more frequently cited financial constraints, fear of the procedure, and interference from personal contacts. No significant gender differences were observed for fear of looking worse.

## Discussion

The study examined clinical and gender-specific patterns of BDD symptoms across a spectrum of severity, focusing on prevalence and perceived outcomes of aesthetic procedures. By comparing individuals meeting full BDD, subclinical BDD, and individuals without meeting formal diagnostic BDD criteria, using validated self-report measures in a large sample, the findings offer valuable insights into appearance-related concerns and cosmetic treatment utilization and attitudes across the BDD symptom continuum.

### Patterns of cosmetic surgery behavior in BDD

The present findings first highlight the prevalence of cosmetic surgery behavior in relation to BDD symptom severity. Approximately 30 % of individuals meeting DSM-5 criteria for BDD reported having undergone at least one cosmetic procedure, with significantly higher rates than those observed in the subclinical and appearance concerns control groups. The rate of planned or desired future surgeries was even higher – particularly among those with a history of prior cosmetic interventions. Surgical targets in the BDD group most frequently involved the nose, face and breast, reflecting patterns found in previous studies that identified facial features as a central focus in BDD [e.g., 34]. In contrast, the subclinical group showed more frequent concerns related to general body dissatisfaction such as stomach and weight, which are less specific to BDD’s core features. Overall, the findings on cosmetic treatment utilization and attitudes complement and confirm earlier studies documenting the increased prevalence of BDD in dermatological and plastic surgery settings [e.g., 35,36].

Cognitive-behavioral models of BDD suggest that while aesthetic procedures may offer temporary relief, they reinforce dysfunctional appearance beliefs and perpetuate symptom maintenance [38]. Accordingly, cosmetic surgeries seldom lead to lasting improvement in core BDD symptoms [3,10]. In line with this, average satisfaction with past surgeries was significantly lower in the BDD group. This finding corresponds with longitudinal findings suggesting that cosmetic procedures rarely resolve core body image concerns [3], often fueling further aesthetic-treatment-seeking behavior. Furthermore, we found that individuals with BDD expressed a substantially stronger desire for future cosmetic procedures compared to subclinical and individuals with appearance concerns, highlighting their pronounced treatment-seeking behavior centered on aesthetic interventions.

Additionally, hierarchical regression showed that higher BDD symptom severity was associated with an increased interest in cosmetic surgery. Including conviction into appearance-related beliefs significantly enhanced this prediction, suggesting that both symptom severity and poor insight independently contribute to the interest in cosmetic procedures. This aligns with previous research identifying poor insight as a core feature of BDD, linked to stronger conviction in distorted appearance beliefs [7,8]. Previous research has shown that insight can fluctuate over time and across situations in individuals with BDD [8], which might be an explanation why its predictive value varies depending on the outcome assessed. While current conviction and current interest in cosmetic surgery are measured simultaneously, past decisions to undergo surgery may be less directly tied to current levels of insight. Moreover, the distinction between cognitive variables such as conviction and actual behavior may also play a role, as intentions do not necessarily translate into actions. Additionally, actual cosmetic procedures could be influenced by a range of contextual factors such as financial resources, access to medical services, or social support, further differentiating them from expressed intentions.

Notably, financial constraints were the most commonly cited barrier to surgery across all groups. However, fear of looking worse following surgery was significantly more prevalent in the BDD group, possibly reflecting the perfectionistic standards, negative interpretation biases and heightened self-focused attention that characterize the disorder [39]. This fear may also indicate a generally negative affective appraisal of one’s appearance and an implicit recognition that cosmetic procedures do not address the core problem, possibly contributing to feelings of hopelessness and perpetuating the cycle of treatment-seeking. Overall, these results reinforce the notion that cosmetic surgery is not an effective long-term solution for BDD symptoms, and that surgery-related behaviors are strongly associated with symptom severity.

Finally, the study underscores that despite increasing awareness of BDD over the past decades, BDD often goes undetected despite its high symptom burden and frequent cosmetic treatment engagement. Only 9.4 % of individuals meeting BDD criteria reported having received a formal diagnosis. This gap likely reflects multiple barriers to mental health care, including stigma, lack of insight, and gender-specific factors that hinder help-seeking and diagnosis [6]. Additionally, 44.5 % of the BDD sample reported at least one prior mental health diagnosis, predominantly affective and anxiety disorders, in line with established comorbidity profiles for BDD [e.g., 39]. Moreover, participants with BDD exhibited elevated depressive symptoms and lower educational attainment, consistent with research documenting the disorder’s negative impact on psychosocial and academic functioning [3]. These findings highlight the clinical need for improved diagnostic screening and psychological evaluation, particularly before undertaking cosmetic procedures.

### Gender differences in BDD presentation

Gender-based comparisons revealed distinct patterns in cosmetic surgery behavior and aesthetic concerns among individuals with BDD. While women and men did not significantly differ in the overall likelihood of having undergone cosmetic surgery, gender-specific patterns emerged in the types of procedures pursued. Women were significantly more likely to have undergone breast surgery, whereas men were more likely to have undergone hair-related procedures, aligning with prior findings that highlight hair loss and muscularity-related concerns as prominent in male BDD presentations [4,15]. Women also demonstrated a significantly higher intention to pursue future procedures – a pattern consistent with literature on internalized appearance norms and heightened aesthetic dissatisfaction among women [41–43]. Notably, they also reported significantly higher overall symptom severity. These gender differences may reflect broader cultural appearance ideals and sociocultural pressures, which are particularly pronounced for women and have been linked to the development and maintenance of body image disturbances [44,45]. However, when interpreting these findings, it should be considered that men may be less likely to openly report psychological distress or appearance-related concerns, which could lead to an underestimation of their symptom severity in self-report measures [46].

Interestingly, within the subgroup who had undergone cosmetic surgery, no significant gender difference in BDD symptom severity was found. This indicates that, among individuals who decide to undergo cosmetic procedures, symptom severity does not differ significantly between genders, despite gender-specific concerns and differences in the types of procedures pursued. Given the limited existing research on gender differences in symptom severity post-cosmetic intervention, this represents an important area for future investigation.

However, gender differences in insight have been less consistently reported. For example, Phillips et al. [14] found no significant gender differences in insight among individuals with BDD. Our finding of higher conviction in women (even after controlling for symptom severity) may reflect gender-specific sociocultural factors, such as stronger internalization of appearance ideals among women [e.g., 46]. Moreover, insight in BDD has been identified as fluctuating, even over the course of a couple of days [8], which may further contribute to variability in findings across studies. Thus, future research should investigate these gender differences in insight more systematically, considering temporal fluctuations and social influences.

Barriers to cosmetic surgery also varied by gender. Women more frequently cited financial concerns, fear of the procedure, and discouragement from others as reasons against cosmetic surgery. These barriers reflect previous research identifying cost as a common deterrent and the stronger influence of social factors, such as vicarious experiences through family or friends, on women’s decisions regarding cosmetic procedures [48]. In contrast, men reported significantly higher full-time employment rates and fewer social or financial deterrents, potentially facilitating access to cosmetic interventions.

Beyond aesthetic surgery utilization and attitude, further gender differences emerged in psychosocial and clinical domains. Interestingly, women were significantly more likely to report being in a romantic relationship, which is consistent with prior research suggesting gender differences in social functioning and suggests that women may be more likely to maintain close interpersonal relationships despite symptom severity [14]. In contrast, men were more likely to report never having received a mental health diagnosis, despite exhibiting clinically relevant symptoms, underscoring gender-specific barriers to mental health recognition and help-seeking [49,50].

These findings emphasize the importance of gender-sensitive approaches in the assessment and treatment of BDD, acknowledging both shared and distinct pathways in the manifestation of the disorder.

### Limitations

Despite the strengths of the present study – including a large sample size and comprehensive assessment of BDD-related behaviors – several limitations must be acknowledged. First, the categorization into the different groups (BDD, subclinical, appearance concerns control) was based on self-report measures rather than structured clinical interviews, which may reduce diagnostic accuracy. However, as described above, the measures used to establish the group status were closely linked to current BDD DSM-5 criteria. Still, while this approach enabled the recruitment of a large and thus potentially more representative sample – unprecedented in terms of sample size within this research area – it limits the clinical validity. Future studies should incorporate clinician-administered interviews to validate self-reported BDD status.

Second, the online recruitment strategy may have introduced selection bias, as individuals experiencing higher levels of distress may have had a different likelihood to participate than those with lower levels of distress. This could limit the generalizability of the findings to the broader BDD population. However, since the majority of the sample met full BDD criteria, this somewhat mitigates concerns about representativeness. Still, the predominantly female, German-speaking sample may restrict applicability to other cultural and demographic groups.

Third, retrospective self-reports on cosmetic procedures, psychological diagnoses, and satisfaction with past surgeries introduce the possibility of recall bias, cognitive distortions, and social desirability effects. This is especially relevant for individuals currently experiencing BDD symptoms, who may evaluate past surgeries more negatively. Moreover, the cross-sectional design of the study precludes causal inferences regarding the relationship between BDD symptom severity, cosmetic interventions, and postoperative satisfaction.

Fourth, the group without a BDD status consisted of individuals who completed a BDD screening despite not endorsing clinically relevant symptoms. It remains unclear what motivated these individuals to take part in a BDD-related study, raising questions about the extent to which this subgroup represents the general population. Future research may benefit from including a comparison group drawn from the broader public without self-selection based on body image concerns.

Finally, the assessment of illness insight was based on a single-item measure, which may not fully capture the complexity of this construct. Future studies should consider more nuanced and validated instruments to examine the role of insight in BDD more comprehensively. Additionally, expanding the investigation of motives and expectations underlying decisions for cosmetic procedures could offer further clarity regarding treatment trajectories and long-term outcomes. Furthermore, the study did not assess non-binary gender identities, limiting insights into gender diversity.

### Clinical implications

This study highlights the substantial clinical burden of BDD, characterized by high symptom severity, diverse appearance concerns, and frequent engagement in cosmetic procedures. In light of the consistently low satisfaction with past cosmetic interventions and the strong desire for future surgeries, comprehensive patient-centered counseling and psychoeducation should be prioritized prior to any aesthetic intervention. Such consultations should also integrate systematic assessment of illness insight, which has been shown to independently predict interest in cosmetic surgery. Addressing these factors through treatment concepts that include specific modules addressing the wish for cosmetic procedures [51] may help to manage expectations and reduce potentially harmful or unnecessary interventions.

The gender-specific pattern of cosmetic surgery interest, particularly the higher intention among women to undergo future procedures, highlights a critical point for tailored prevention and early intervention strategies. In particular, integrated care models addressing both BDD symptoms and common psychological comorbidities, such as mood and anxiety disorders, are essential to effectively meet the diverse clinical needs and improve patient outcomes.

Moreover, the substantial under-recognition of BDD within this population calls for enhanced screening protocols in cosmetic and dermatological settings. Training for primary care providers, dermatologists, and cosmetic surgeons on BDD identification and referral pathways is critical to facilitate early diagnosis and appropriate mental health care. Additionally, efforts to reduce patient-related treatment barriers are essential to improve help-seeking behavior and engagement in mental health services.

## Conclusion

This study offers one of the most comprehensive assessments to date of BDD cosmetic treatment utilization and attitudes and gender-specific patterns in a large sample. By systematically examining individuals across a continuum from appearance concerns to subclinical and full BDD, this study sheds light on differences in cosmetic surgery engagement and satisfaction, highlighting patterns that may be less apparent in clinical samples. Importantly, the findings confirm that cosmetic procedures rarely alleviate core BDD symptoms and may contribute to perpetuating dysfunctional appearance beliefs and ongoing cosmetic treatment-seeking behavior. Beyond symptom severity, the role of insight emerges as a critical factor influencing cosmetic surgery intentions, underscoring the importance of integrating cognitive assessments into clinical practice. The clear gender differences in symptom severity, treatment interest, and psychosocial factors highlight the need for tailored, gender-informed interventions.

Future research should expand this work by including gender-diverse populations and further investigating psychological mechanisms such as insight and motivation underlying cosmetic decision-making, to better inform prevention and treatment strategies.

## Data Availability

All relevant data are within the manuscript and its Supporting Information files. This is because we do not have ethics approval to upload the entire data set.

## Supporting information

**S1 Table. Demographic and clinical characteristics of participants with self-reported BDD.**

**S2 Table. Comparisons of BDD status groups.**

**S3 Table. Comparisons of female and male individuals with a diagnosis of BDD**

## Notes

### Competing Interest Statement

The authors have declared no competing interest.

### Funding Statement

The author(s) received no specific funding for this work.

### Author Declarations

The study was approved by the Ethics Committee of Faculty 7 at the University of Münster (Approval No. 2017-32-JSch). Participants were informed about the voluntary nature of participation, confidentiality, and the use of their data for research purposes. Informed consent was obtained digitally at the beginning of the online survey, and participants were provided the option to withdraw at any time without consequences.

